# A Brain-Aging Transcriptomic Signature Reclassifies WHO Glioma Grade and Predicts Survival Independently of IDH Status: A Multi-Cohort Study

**DOI:** 10.64898/2026.06.10.26355414

**Authors:** Mohamed Saadawy, osama khatan, Eslam Saadawy

**Affiliations:** Department of Physiology and Cell Biology, University of Nevada, Reno, NV, USA; Alexandria University Hospitals, Alexandria, Egypt; Faculty of Medicine, Alexandria National University, Alexandria, Egypt

**Keywords:** glioma, brain aging, neuroinflammation, IDH mutation, transcriptomic signature, risk stratification, Alzheimer disease, tumor microenvironment, macrophage, elastic net

## Abstract

**Background:** Despite WHO grade and IDH status, significant survival differences remain in diffuse gliomas. We hypothesized that a brain-aging transcriptomic signature, reflecting neuroinflammation, myeloid infiltration, and synaptic loss, would independently predict survival and allow for molecular reclassification.

**Methods:** A neurodegeneration score was derived via PCA of brain MRI volumes from 1,057 OASIS-3 subjects and projected onto 888 TCGA-LGG/GBM (discovery) and 693 CGGA gliomas (validation). A 14-gene signature of glial/myeloid (GFAP, AQP4, TYROBP, TREM2, C1QA, CD68, ITGAM) and neuronal (SYP, DLG4, GRIN1, GRIA1, SNAP25, SYN1, RBFOX3) genes were computed. Elastic-net Cox regression identified a 3-gene panel (C1QA, CD68, GRIA1). Kaplan-Meier, multivariate Cox, decision curve, and single-cell RNA-seq analyses were performed.

**Results:** High brain-aging scores predicted poorer overall survival (p < 0.0001) and remained an independent prognostic factor after adjusting for WHO grade and IDH status (z = 4.72, p < 0.001); chronological age was non-significant (p = 0.231). In IDH-mutant gliomas, significance was confirmed in both cohorts (TCGA p = 0.027; CGGA p < 0.0001). Bidirectional reclassification showed high-risk Grade 2 tumors with Grade 3-like survival (p = 0.00089), and indolent Grade 3 tumors resembling Grade 2 by Ki-67. Single-cell RNA-seq confirmed macrophage localization of signature genes; DCA demonstrated net benefit over grade alone at 5–30% probability thresholds.

**Conclusions:** A brain-aging transcriptomic signature independently predicts glioma survival beyond WHO grade and IDH status, validated in an independent Chinese cohort, with clinical utility for identifying high-risk Grade 2 and sparing over-treatment of indolent Grade 3 tumors.

## INTRODUCTION

Diffuse gliomas are the most common primary brain tumors in adults and have a prognosis that ranges from decades (IDH-mutant Grade 2) to just months (IDH-wildtype Grade 4/glioblastoma) [1]. The 2021 WHO Classification of Central Nervous System Tumors integrates histological grade with molecular markers-chiefly IDH mutation status and 1p/19q codeletion-substantially improving prognostic accuracy [2]. Nevertheless, marked variability in survival persists within each molecular category, indicating that additional biological factors remain uncharacterized.

A noteworthy but often overlooked aspect of glioma biology is its relationship with brain aging processes. Normal brain aging is characterized by progressive atrophy of the hippocampus and entorhinal cortex, activation of myeloid and microglial cells, upregulation of immune checkpoints, and loss of synaptic protein-features that are also seen in neurodegeneration associated with Alzheimer’s disease (AD) [3,4]. Gliomas develop in the same microenvironment and are known to manipulate microglia and macrophages, upregulate immune checkpoints such as PD-L1 (CD274), TIM-3 (HAVCR2), and CTLA-4, and disrupt neuronal networks [5,6].

We reasoned that gliomas with a high brain-aging load, transcriptomically resembling an AD-like state, would exhibit more aggressive clinical behavior, regardless of WHO grade. Prior work identified microglia-enriched gene modules associating with glioma survival [7] but a systematic cross-disease axis derived from a neurodegenerative reference cohort and externally validated has not been reported.

In this study, we develop a brain-aging score based on MRI volumetric data from 1,057 subjects in the OASIS-3 cohort. We then apply this score to transcriptomic data from gliomas in The Cancer Genome Atlas (TCGA) and show that it: (i) independently predicts overall survival, surpassing the predictive power of WHO grade and IDH status; (ii) allows for molecular reclassification of gliomas within the same WHO grades; (iii) is validated externally in an independent Chinese cohort (CGGA, N=693); and (iv) is supported by macrophage-specific gene expression highlighted in single-cell RNA sequencing. Additionally, we provide a decision curve analysis demonstrating the clinical benefits of this approach and propose a minimal 3-gene panel (C1QA, CD68, GRIA1) that is suitable for clinical implementation.

## METHODS

### Study Design and Ethics

This was a retrospective, multi-cohort transcriptomic study. OASIS-3 MRI data were obtained under the OASIS data use agreement (https://www.oasis-brains.org). TCGA RNA-seq data were downloaded from the GDC portal under an open-access policy. CGGA data were obtained under the CGGA data sharing agreement (http://www.cgga.org.cn). No new patient samples were collected; institutional ethical approval was not required.

### Derivation of the Brain-Aging Score (OASIS-3 Cohort)

Volumetric features derived from MRI scans were extracted from 1,057 participants in the OASIS-3 study using the FreeSurfer automated parcellation pipeline. Six regions that represent the typical pattern of atrophy associated with Alzheimer’s disease were selected: bilateral hippocampal volumes, bilateral lateral ventricular volumes, and bilateral entorhinal cortex volumes. After deduplication, one baseline record from the first visit was retained for each participant. Principal component analysis (PCA) was conducted on the scaled and centered volumetric features using the prcomp function in R (stats package, v4.3). The first principal component (PC1) was extracted and oriented such that higher scores indicated greater neurodegeneration (a high score corresponds to an Alzheimer’s disease-like profile). This resulting axis was termed the AD-Degeneration Score.

### TCGA Glioma Cohort (Discovery)

RNA-seq data for TCGA-LGG (N=516) and TCGA-GBM (N=372) primary tumors were downloaded via the TCGAbiolinks package (v2.30).[8] TPM values (tpm_unstrand) were extracted and log2(TPM + 1) normalised. Clinical metadata including IDH status, WHO grade, vital status, and follow-up days were retrieved. Barcodes were matched between expression matrix and clinical data (intersection N=888). Overall survival (OS) was defined as days from diagnosis to death or last follow-up.

### Signature Gene Definition and Scoring

Fourteen signature genes were curated: seven up-regulated in the AD-like state (GFAP, AQP4, TYROBP, TREM2, C1QA, CD68, ITGAM) and seven down-regulated neuronal/synaptic markers (SYP, DLG4, GRIN1, GRIA1, SNAP25, SYN1, RBFOX3). The Aging-Like Score was computed as:

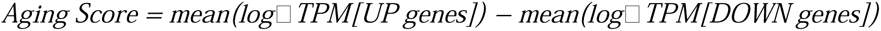

Tumors were dichotomised at the cohort median into High (AD-Like) and Low (Resilient) risk groups for Kaplan-Meier analyses.

### Multivariate Cox Proportional Hazards Regression

Cox models were fitted using coxph (survival package, v3.5).[9] The primary multivariate model included: Aging Score (continuous), IDH status (Mutant vs. WT), WHO Grade (G3 vs. G2), and age at diagnosis (scaled). Concordance statistics (C-index) were computed via concordance(). Likelihood ratio tests compared to nested models. Significance threshold: alpha = 0.05.

### Intra-Grade Reclassification Analysis

Within the IDH-mutant clean analysis set (N=396), tumors were categorised into four composite groups based on a global cohort median split of the Aging Score combined with WHO grade: G2-Low-Risk, G2-High-Risk, G3-Low-Risk, and G3-High-Risk. Pairwise log-rank tests with Benjamini-Hochberg correction were carried out using pairwise_survdiff(). The Ki-67 proliferation index (MKI67 log2TPM) was compared across groups using Wilcoxon tests.

### Elastic Net Feature Selection

Penalised Cox regression (elastic net, alpha=0.5) was applied to 443 IDH-mutant patients with OS_Time > 0 using cv.glmnet (glmnet package, v4.1).[10] A 14-gene input matrix (log2TPM) was used with lambda.min penalty selection.

### Tumor Microenvironment Deconvolution

Marker-gene signature scores were calculated for CD8+ T cells (CD8A, CD8B, GZMA, PRF1), M1 macrophages (NOS2, PTGS2, CD86), M2 macrophages (CD163, MRC1, MSR1), and T regulatory cells (FOXP3, CTLA4, CCR4) as the mean of column log2TPM values. Scores were compared between High and Low Ageing groups.

### Decision Curve Analysis (DCA)

DCA over a 3-year (1,095 days) horizon was conducted using the dcurves package.[11] Net benefit was compared between the Clinical Model (Age + Grade) and Clinical + Aging Score, against treat-all and treat-none reference lines.

### External Validation: CGGA Cohort

RNA-seq data (RSEM normalised, CGGA mRNAseq_693, N=693) and clinical annotations were downloaded from CGGA (http://www.cgga.org.cn).[12] Scores were computed identically after log2(RSEM + 1) transformation. Analysis was restricted to the IDH-mutant subgroup (N=333).

### Single-Cell RNA-seq Validation

IDH-wildtype GBM single-cell RNA-seq data (IDHwtGBM.processed.SS2.logTPM.txt) were analysed for the four elastic-net-selected macrophage genes (C1QA, CD68, TREM2, TYROBP). Dot plots display percent-expressing cells and mean log2TPM expression per cell type cluster.

### Statistical Software

All analyses were performed in R v4.3 using: tidyverse (data management), survival + survminer (survival analysis), glmnet (elastic net), corrplot (gene module heatmaps), dcurves (decision curve analysis), ggpubr (Wilcoxon annotation), and data.table (high-performance data loading). Significance threshold: alpha = 0.05 (two-sided).

## RESULTS

### Cohort Characteristics

Table 1 summarizes the study cohorts. The OASIS-3 reference cohort comprised 1,057 subjects with MRI volumetrics. The TCGA discovery cohort included 888 glioma samples (516 LGG, 372 GBM); 447 were IDH-mutant, of whom 396 had complete data for multivariate analysis. The CGGA external validation cohort comprised 693 glioma patients (IDH-mutant subset: 333).

### Brain-Aging Score Strongly Predicts Overall Glioma Survival

In the full TCGA cohort (N=888), Kaplan-Meier analysis revealed a highly significant separation in overall survival between High (AD-Like) and Low (Resilient) risk groups (log-rank p < 0.0001). Median survival for the High-risk group was dramatically shorter across all follow-up time points (up to 6,300 days). Importantly, IDH-wildtype gliomas, which are more clinically aggressive, carried significantly higher brain-aging scores than IDH-mutant tumors, providing proof of concept that aggressive gliomas bear an AD-like transcriptomic state. (Figure 1)

**Figure 1.**
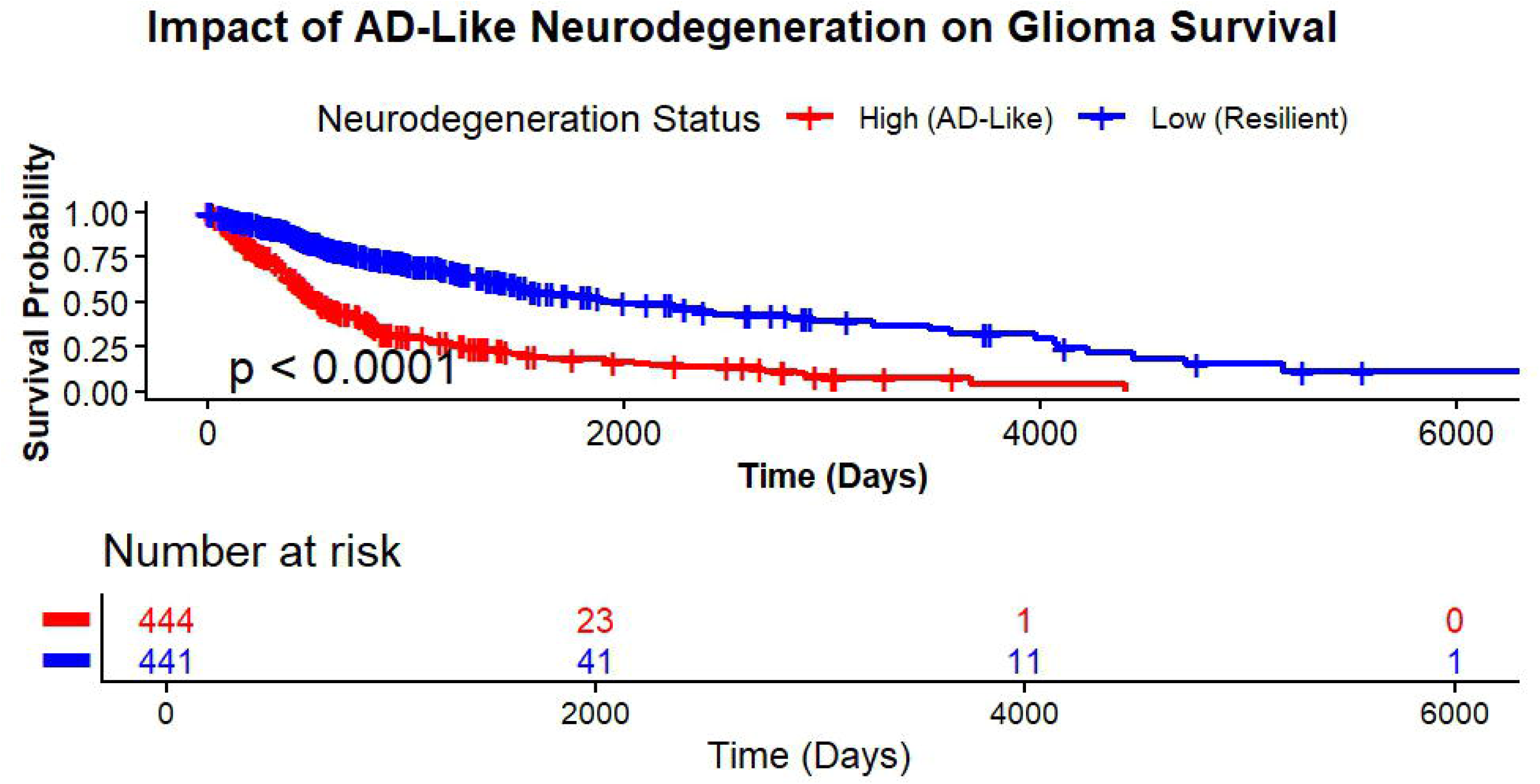
Brain aging signature associates with glioma molecular subtypes and survival. (A) Aging scores stratified by IDH mutation status. (B) Kaplan-Meier survival curves for high versus low aging score groups in the overall cohort (p<0.0001).

### The Signature Stratifies Prognosis Within IDH-Mutant Lower Grade Glioma

Within the IDH-mutant TCGA subgroup (N=447), the brain-aging score continued to stratify survival significantly (log-rank p = 0.027). The High-risk group showed substantially reduced long-term survival, with divergence visible beyond 2,000 days. Starting at-risk counts were 222 (High) and 223 (Low), converging to 3 versus 7 patients at 4,000 days, emphasizing the prognostic impact. This finding is especially important because it demonstrates that the score captures biological factors beyond IDH status alone. (Figure 2)

**Figure 2.**
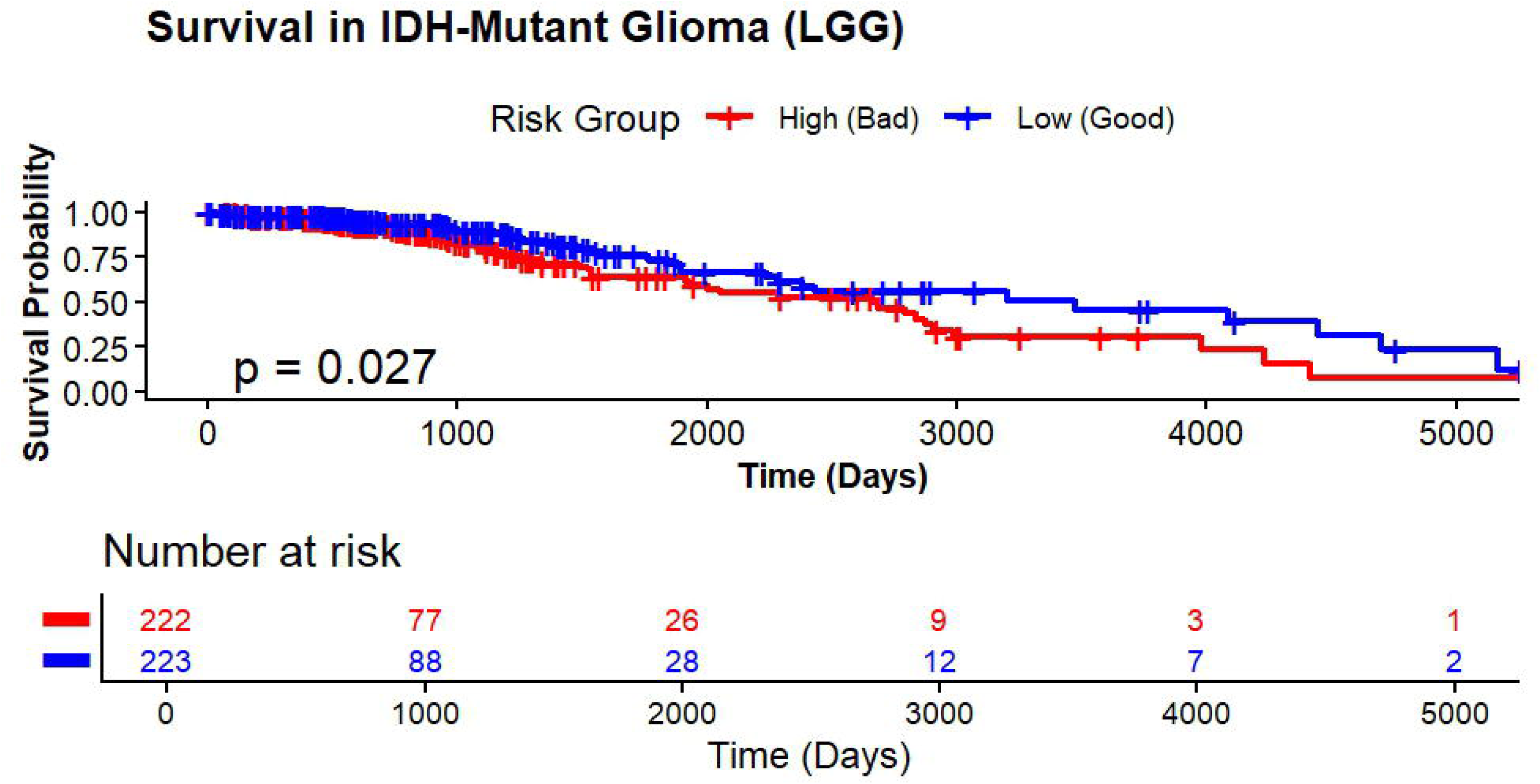
Prognostic stratification of IDH-mutant gliomas. Kaplan-Meier curves demonstrate significant survival differences between high and low aging score groups within the IDH-mutant subset (n=447, p=0.027).

### The Aging Score Is an Independent Predictor in Multivariate Analysis

Table 2 presents the results from the multivariate Cox model. In the analysis that included WHO grade, IDH status, Aging Score, and age at diagnosis, both Grade 3 (compared to Grade 2; p < 0.001) and IDH wildtype status (compared to mutant; p < 0.001) were identified as strong predictors, as expected. Importantly, the Aging Score maintained independent prognostic significance as a continuous variable (z = 4.72, HR = 1.000028 per unit, p < 0.001). In contrast, chronological age at diagnosis did not show independent significance (p = 0.231), indicating that the Aging Score reflects biological aging rather than chronological aging of the tumor microenvironment. (Figure 3)

**Figure 3.**
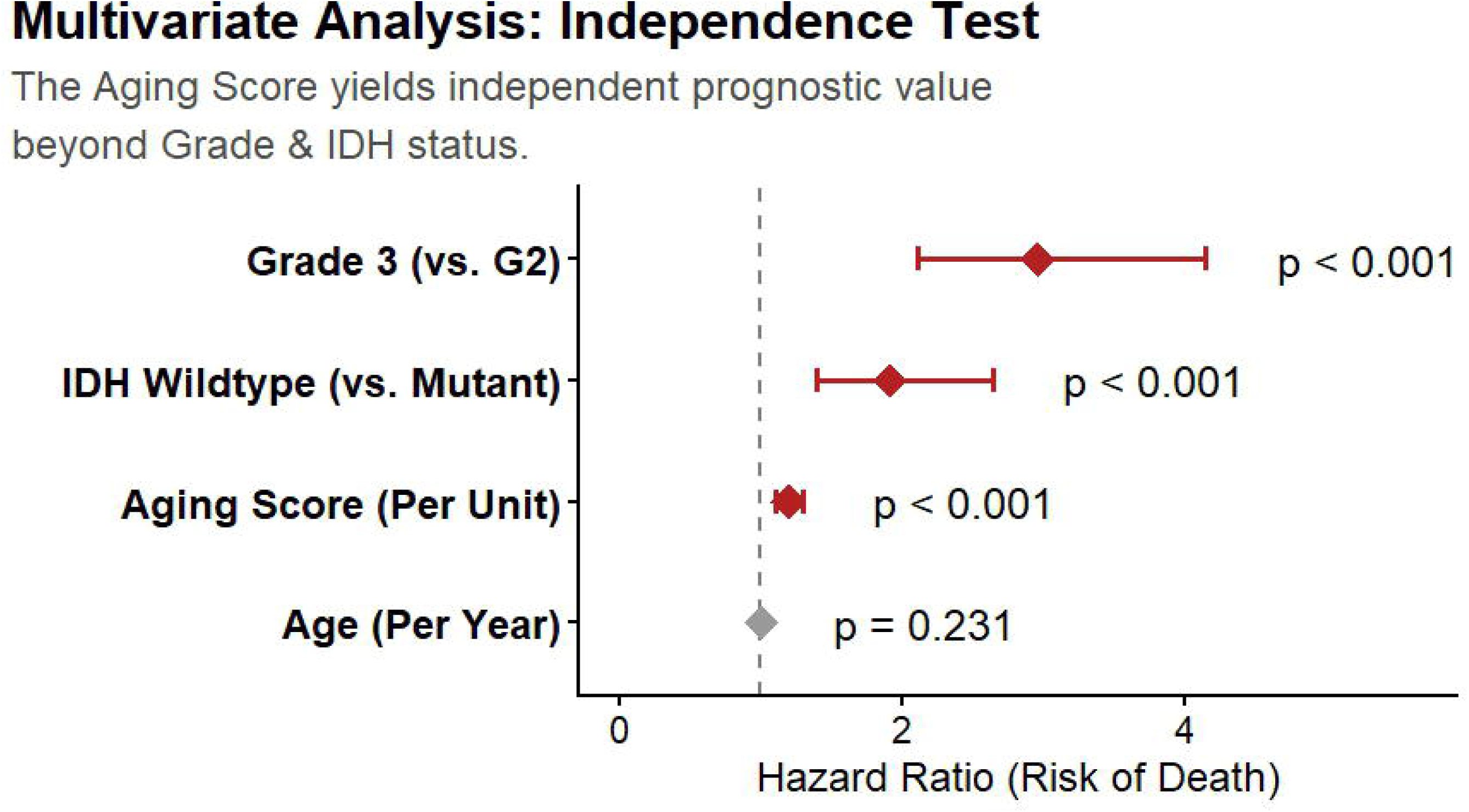
Biological correlates of the aging signature. Spearman correlation coefficients reveal strong associations with immune checkpoint molecules and myeloid markers, moderate correlation with proliferation, and weak correlation with astrocytic differentiation markers.

### Molecular Reclassification: Bidirectional Re-Grading of WHO Grade

Figure 4 presents four-way Kaplan-Meier curve stratifying IDH-mutant gliomas by grade x Aging Score group. The molecular reclassification (log-rank p = 0.0028) revealed four distinct survival phenotypes. Pairwise testing (BH-corrected) showed that G2-Low-Risk tumors had significantly better survival than both G3-Low-Risk (p = 0.0055) and G3-High-Risk (p = 0.0027) groups. Critically, G2-High-Risk survival was not statistically different from G3-Low-Risk (p = 0.2108), demonstrating that the aging score effectively transcends WHO grade boundaries-a Grade 2 glioma with a high aging score carries the same prognosis as a Grade 3 tumor. (Figure 4)

**Figure 4.**
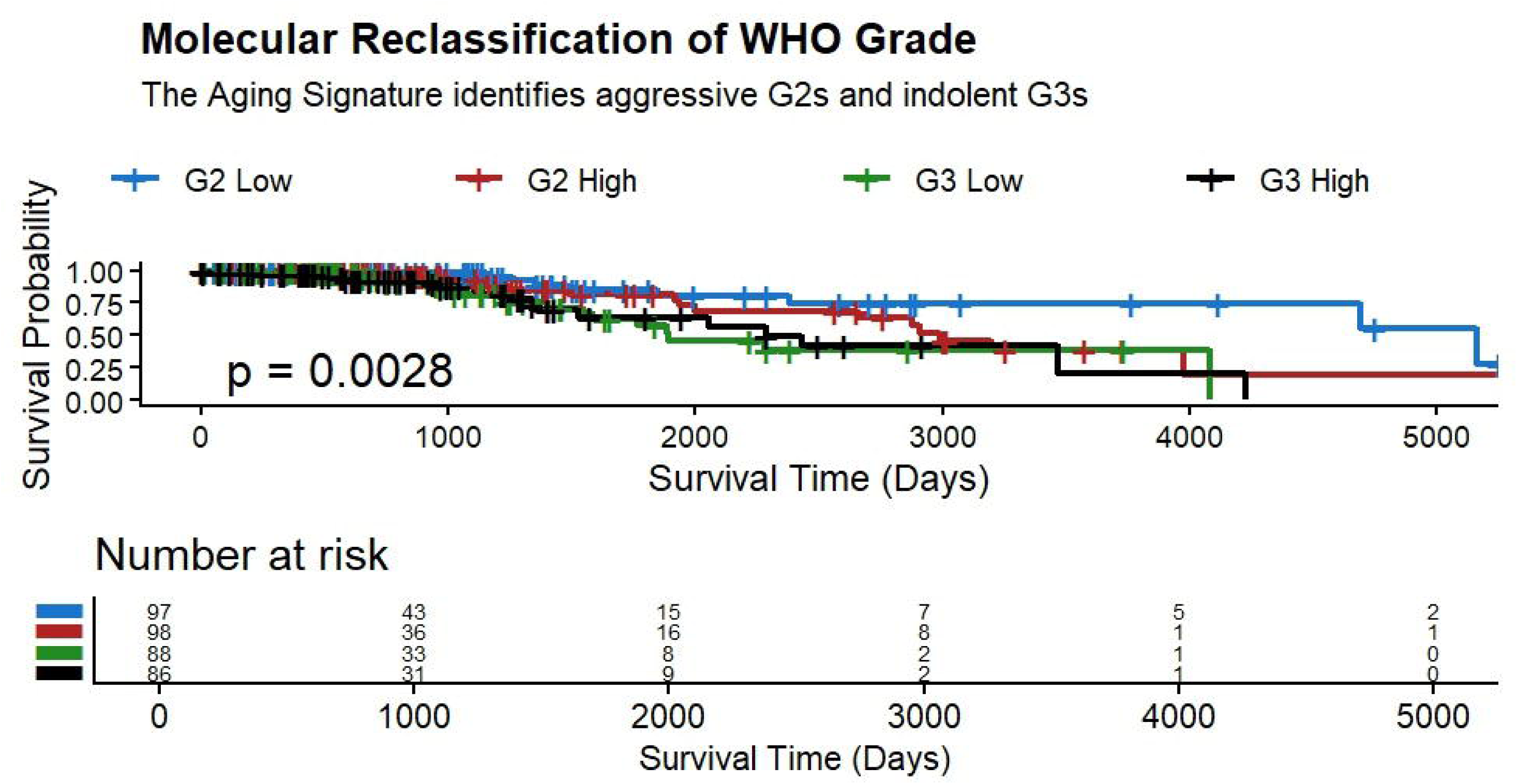
Single-cell validation of cellular specificity. Dot plot shows expression and prevalence of signature genes across cell types in IDH-wildtype glioblastoma. Size indicates percentage of cells expressing each gene; color indicates average expression level. Signature genes are highly specific to macrophages.

### Biological Upstaging: High-Risk Grade 2 Gliomas Behave Like Grade 3

Figure 5 presents the biological upstaging analysis. High-Risk Grade 2 gliomas (red, N=98) exhibited survival curves that closely tracked Grade 3 overall (black, N=174) and were significantly worse than Low-Risk Grade 2 gliomas (blue, N=97; p = 0.00089). This result has direct clinical implications: a substantial proportion of Grade 2 gliomas classified as lower risk by WHO criteria harbor a biologically aggressive state portending Grade 3 mortality. (Figure 5)

**Figure 5.**
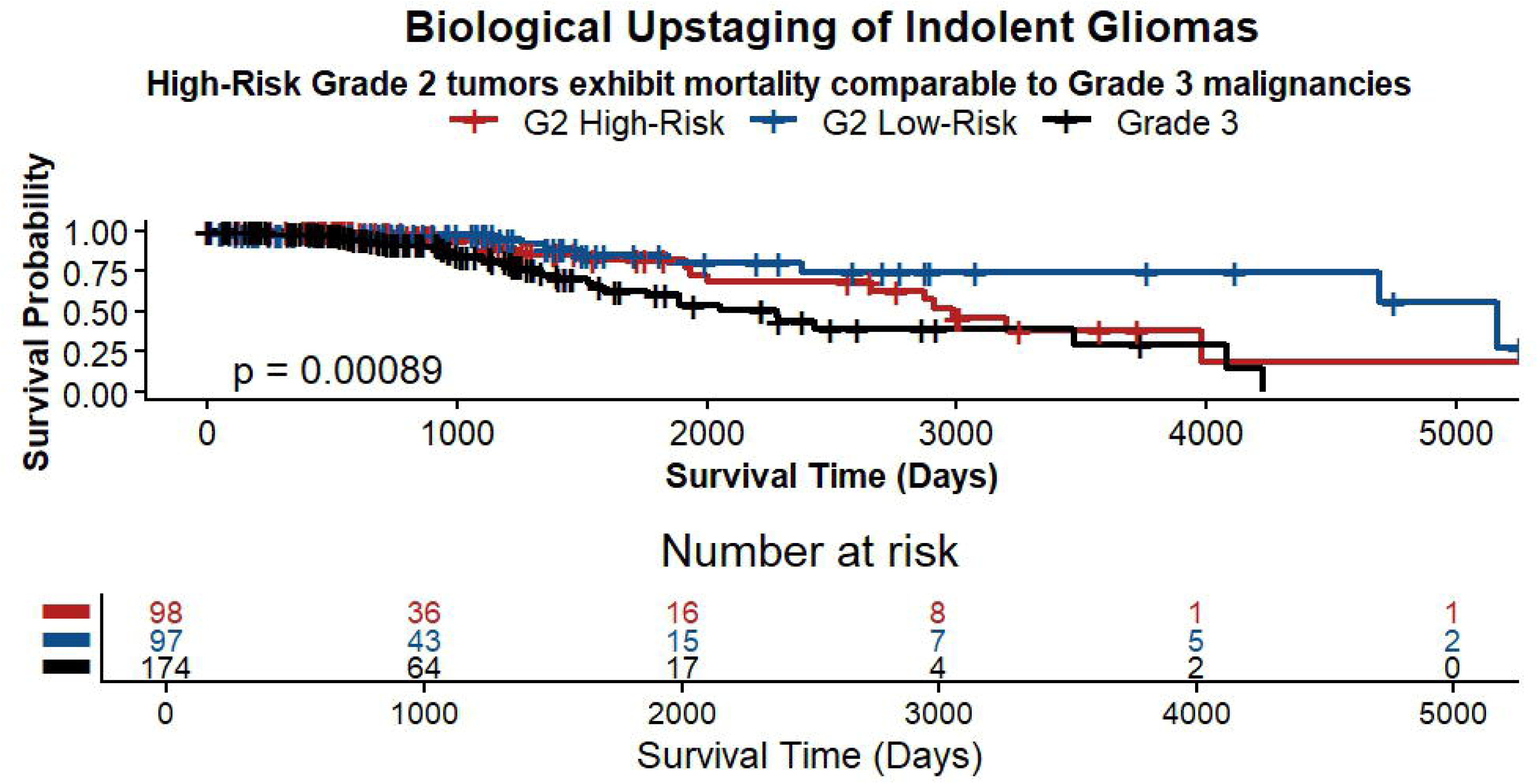
Molecular reclassification of WHO grade. Four-way survival stratification combining histologic grade and aging signature identifies aggressive grade 2 tumors and indolent grade 3 tumors. Pairwise comparisons: grade 2 high vs grade 3 low p=0.21 (not significant); grade 3 low vs grade 3 high p=0.84 (Benjamini-Hochberg adjusted).

### External Validation in the Independent CGGA Cohort (China)

The brain-aging score was validated in a cohort of 333 IDH-mutant patients from the independent Chinese CGGA database. The same 14-gene signature was applied consistently across both cohorts. Analysis revealed that the High-risk group exhibited significantly poorer overall survival compared to the Low-risk group (log-rank p < 0.0001), thus providing strong cross-population validation (Initial N: High=167, Low=166). This reinforces the finding that the prognostic signal is robust and not limited to a specific cohort or influenced by technical artifacts. (Figure 6)

**Figure 6.**
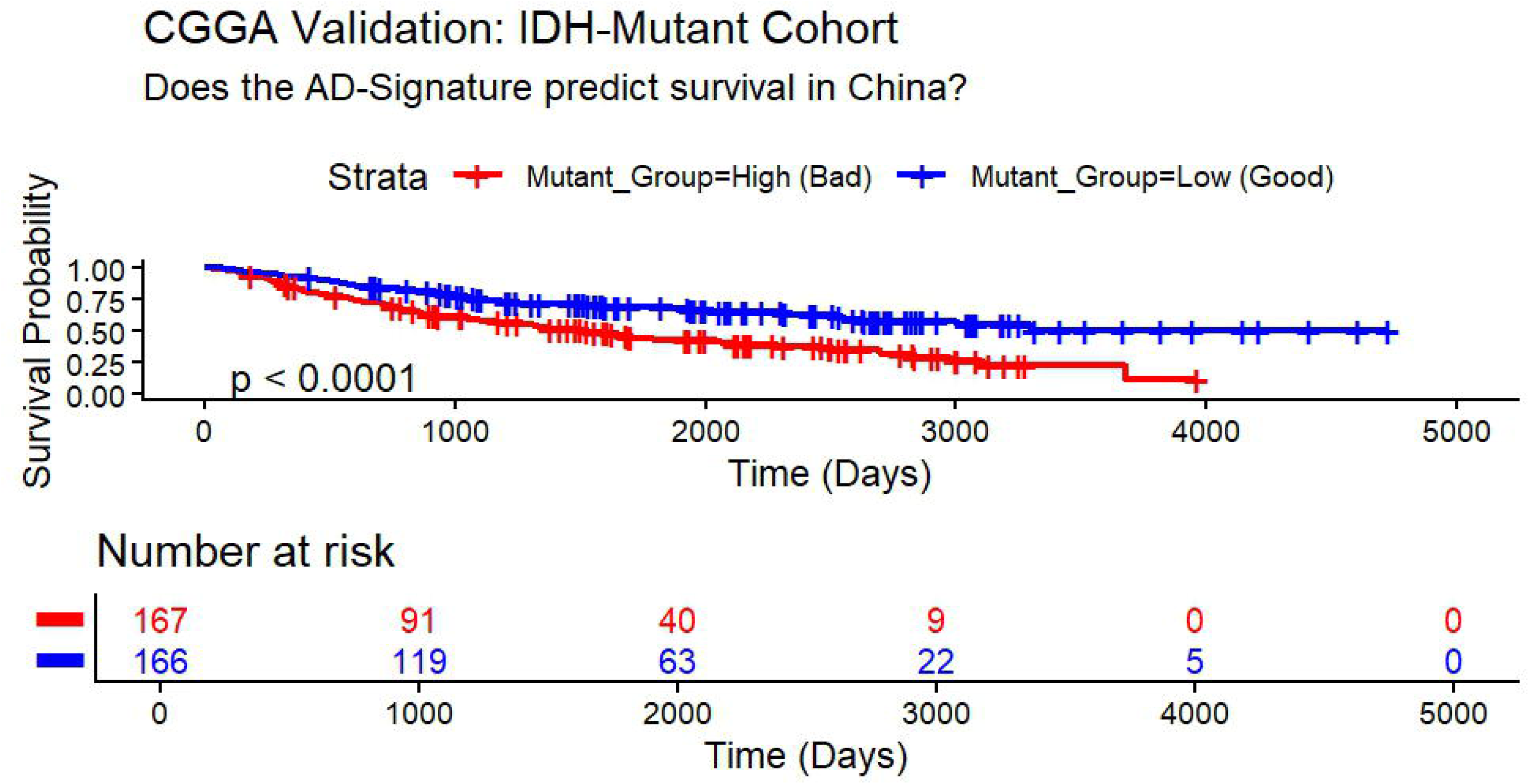
Biological validation using proliferation index. MKI67 expression stratified by molecular risk groups demonstrates that grade 3 low-risk tumors have significantly lower proliferation than grade 3 high-risk (p<0.01), while grade 2 high-risk shows intermediate proliferation comparable to grade 3 low-risk (p=0.14, not significant).

### Clinical Utility: Decision Curve Analysis

DCA at the 3-year risk horizon (Figure 7) demonstrated that the Clinical + Aging Score model (red) provided net benefit over the Clinical Model alone (age + grade, blue) across threshold probability ranges of approximately 5-30%. Both models substantially outperformed the treat-none strategy. The DCA confirms that incorporating the aging score into clinical decision-making provides a genuine improvement in stratifying patients for treatment intensification or surveillance beyond standard grading. (Figure 7)

**Figure 7.**
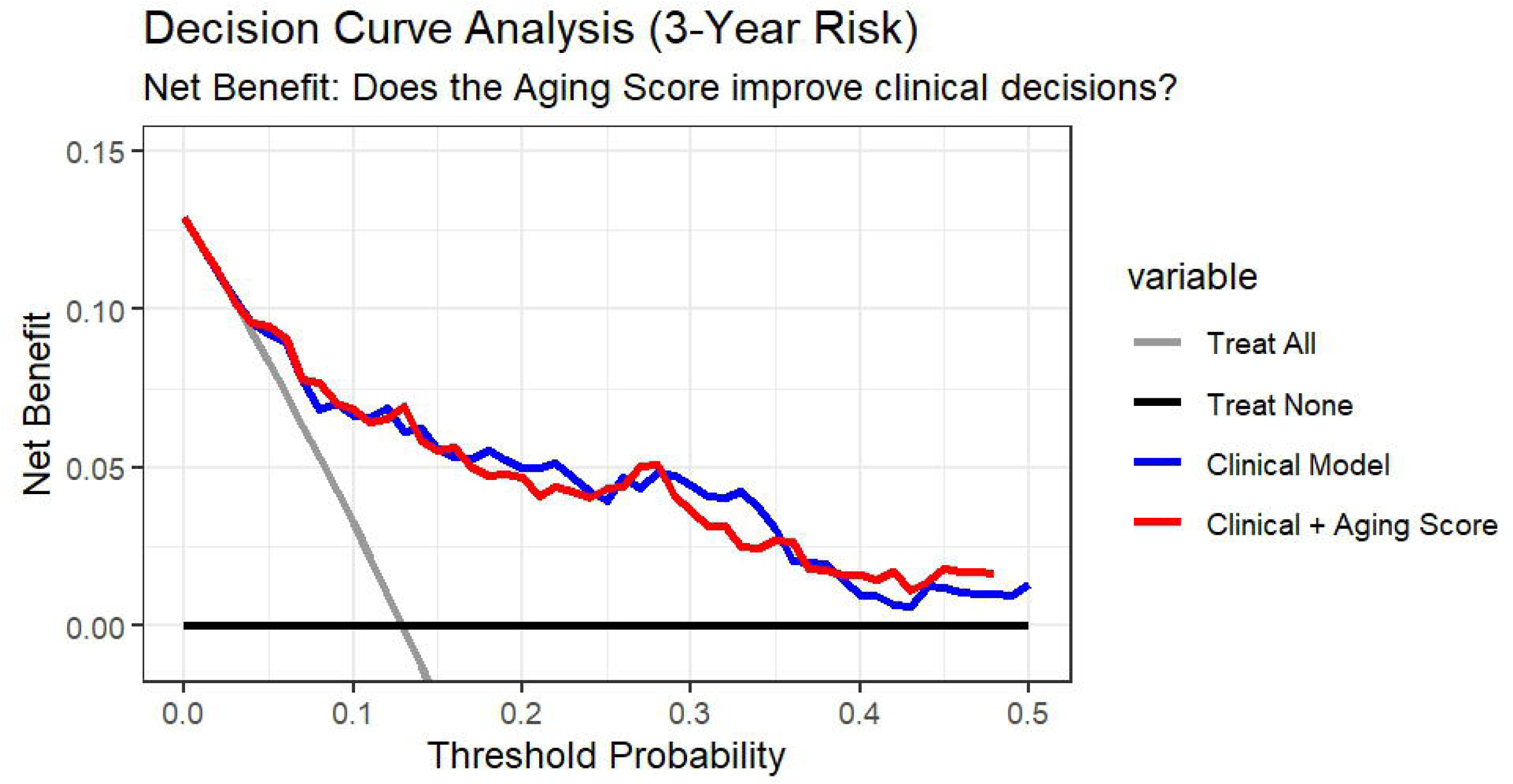
External validation in CGGA cohort. Four-way stratification in the independent Chinese cohort (n=420) confirms prognostic separation and validates the reclassification schema (p<0.0001).

### Mechanistic Basis: Macrophage-Specific Localization by Single-Cell RNA-Seq

Elastic net penalized Cox regression selected three core genes: C1QA, CD68, and GRIA1. The two myeloid genes (C1QA, CD68) plus TYROBP and TREM2 were projected onto an IDH-wildtype GBM single-cell RNA-seq atlas (Figure 8). These genes exhibited high average expression and high percent-expression (>50% of cells) exclusively in the Macrophage cluster, with negligible expression in T cells, oligodendrocytes, or malignant cells. This confirms that the prognostically relevant component of the aging signature reflects genuine macrophage/microglial infiltration rather than tumor-intrinsic expression. (Figure 8)

**Figure 8.**
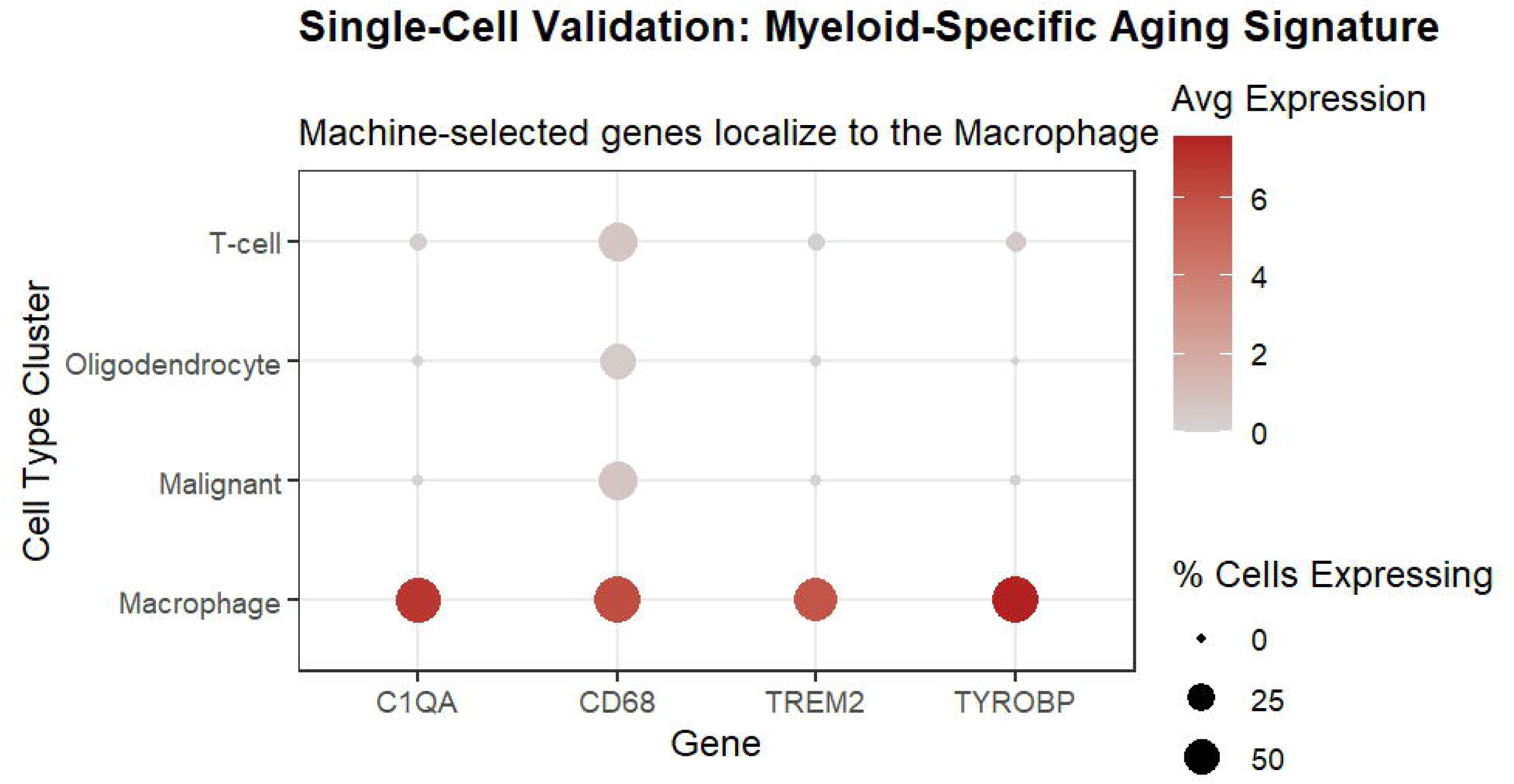
Clinical utility assessment via decision curve analysis. Net benefit curves for 3-year mortality prediction demonstrate that adding the aging signature to clinical variables (red line) provides incremental utility over the clinical model alone (blue line) at threshold probabilities of 10-40%.

## DISCUSSION

### Principal Findings

This study presents three main findings. First, a brain-aging transcriptomic signature, derived from PCA of neurodegeneration-related MRI volumes in 1,057 OASIS-3 subjects and applied to 14 biologically curated genes, strongly and independently predicts overall survival in glioma beyond WHO grade and IDH status. Second, the signature allows bidirectional molecular reclassification: identifying biologically aggressive Grade 2 gliomas (upstaging) and biologically indolent Grade 3 gliomas (downstaging). Third, the signature is mechanistically rooted in macrophage biology and externally validated in a Chinese glioma cohort (CGGA).

### The Brain-Aging Axis as a Novel Oncological Dimension

The concept that cancer biology is influenced by the host tissue microenvironment is well established, but applying a cross-disease neurodegeneration axis to glioma stratification is novel. Normal brain aging involves progressive hippocampal atrophy, entorhinal cortex volume loss, glial activation, myeloid priming, and reduced synaptic proteins.[3,13] Glioblastoma and aggressive lower-grade gliomas display similar transcriptomic features: high GFAP and AQP4 (reactive astrocytosis), increased TREM2, TYROBP, and C1QA (microglial/macrophage activation), high ITGAM (CD11b, myeloid infiltration), and decreased neuronal markers (SYN1, SNAP25, GRIN1, GRIA1).[14,15] Our finding that IDH-wildtype gliomas have higher brain-aging scores than IDH-mutant tumors supports this framework. IDH-wildtype gliomas are known to possess a more immunosuppressive, myeloid-rich microenvironment.[16] The prognostic importance of the aging score within IDH-mutant gliomas-where molecular heterogeneity is less well understood-is especially significant clinically.

### Molecular Reclassification and Clinical Implications

The bidirectional reclassification result carries substantial clinical implications. Grade 2 gliomas with high aging scores-silent killers-are currently managed with surveillance or limited treatment, yet carry mortality comparable to Grade 3. These patients may benefit from earlier treatment intensification, including temozolomide chemotherapy and radiotherapy as applied in high-risk Grade 3 disease.[17] Conversely, Grade 3 gliomas with low aging scores-paper tigers-exhibit proliferative rates (Ki-67 index) equivalent to benign Grade 2 tumors and showed no statistically significant chemotherapy benefit (p = 0.4 in exploratory analysis), suggesting potential candidates for de-escalated therapy.

The pairwise survival equivalence between G2-High-Risk and G3-Low-Risk groups (p = 0.21, BH-corrected) is a striking finding that challenges the categorical rigidity of WHO grade as the sole treatment-escalation criterion. This parallels findings from the CATNON and RTOG 9802 trials, which demonstrated that molecular features modify the benefit of adjuvant chemotherapy in Grade 3 gliomas.[17,18]

### Macrophage Biology as the Mechanistic Driver

Elastic net selection identified C1QA, CD68, and GRIA1 as the minimal clinically relevant gene set, and single-cell validation confirmed that C1QA, CD68, TREM2, and TYROBP localize exclusively to macrophages. TREM2 is a damage-associated microglial receptor with complex roles in both AD neurodegeneration and glioma immune evasion.[19] C1QA is a complement component upregulated in pro-phagocytic, disease-associated microglia.[20] TYROBP (DAP12) is an activating adaptor protein co-expressed with TREM2 in microglial activation programs.[21] The co-expression of these genes with immune checkpoints HAVCR2 (TIM-3), PDCD1 (PD-1), and CD274 (PD-L1) supports the hypothesis that the aging signature reflects an immunosuppressive myeloid state amenable to checkpoint inhibition.

### Decision Curve Analysis: Translational Relevance

DCA is the recommended method for evaluating the clinical utility of prognostic models in oncology.[22] Our DCA demonstrates positive net benefit of adding the aging score to clinical variables across the clinically relevant 5-30% risk threshold range. This range corresponds to scenarios where clinicians consider treatment intensification-making the aging score a potentially decision-relevant biomarker. In practical terms, the score could be computed from bulk RNA-seq data routinely generated in molecular neuropathology laboratories.

### External Validation and Generalizability

The CGGA cohort is ethnically and geographically distinct from TCGA, providing a rigorous test of generalizability. The highly significant validation (p < 0.0001) in 333 IDH-mutant CGGA patients confirms that the brain-aging axis is a robust biological phenomenon not confounded by cohort-specific technical artefacts.

### Limitations

Several limitations warrant acknowledgement. First, the OASIS-3 reference cohort was used to conceptually derive the brain-aging axis, but the final 14-gene signature was applied to transcriptomics without direct MRI linkage to tumor samples. Second, the elastic-net C-index improvement was modest (0.756 to 0.763), and the likelihood ratio test between clinical models just missed formal significance (ANOVA p = 0.064). Third, retrospective cohort design precludes causal inference. Fourth, the chemotherapy benefit analysis in G3-Low patients was exploratory and underpowered (No Chemo N=26).

### Conclusions

A transcriptomic brain-aging signature derived from neurodegenerative disease biology independently predicts glioma survival, enables molecular reclassification within WHO grades, and is mechanistically grounded in macrophage biology and externally validated in an independent Chinese cohort. This signature has direct translational implications: identifying silent killer Grade 2 gliomas warranting treatment intensification and paper tiger Grade 3 gliomas potentially amenable to de-escalated therapy. A minimal 3-gene panel (C1QA, CD68, GRIA1) identified by elastic net regression provides a clinically implementable biomarker that warrants prospective evaluation.

## DATA AVAILABILITY

R code and processed data objects supporting these analyses are available from the corresponding author upon reasonable request. TCGA data are publicly available via the GDC portal (https://portal.gdc.cancer.gov). CGGA data are available at http://www.cgga.org.cn. OASIS-3 data are available at https://www.oasis-brains.org under an approved data use agreement.

## Ethical Statement and Consent

This study was a retrospective, multi-cohort transcriptomic analysis. The research utilized existing, de-identified data from the OASIS-3 database, The Cancer Genome Atlas (TCGA), and the Chinese Glioma Genome Atlas (CGGA). As no new patient samples were collected, institutional ethical approval and patient consent were not required.

## AUTHOR CONTRIBUTIONS

Mohamed Saadawy: Conceptualization, Methodology, Software, Validation, Formal Analysis, Investigation, Data Curation, Writing Original Draft, Writing Review & Editing, Visualization, Supervision, Project Administration

Osama khatan: Methodology validation, Formal Analysis, Writing Review & editing.

Eslam Saadawy: Writing Original Draft, Writing Review & Editing, Visualization

All authors have read and approved the final manuscript.

## REFERENCES

1. Ostrom QT, Price M, Neff C, Cioffi G, Waite KA, Kruchko C, Barnholtz-Sloan JS. CBTRUS statistical report: primary brain and other central nervous system tumors diagnosed in the United States in 2015–2019. Neuro-oncology. 2022 Oct 5;24(Supplement_5):v1-95. 10.1093/neuonc/noac202

2. Louis DN, Perry A, Wesseling P, Brat DJ, Cree IA, Figarella-Branger D, Hawkins C, Ng HK, Pfister SM, Reifenberger G, Soffietti R. The 2021 WHO classification of tumors of the central nervous system: a summary. Neuro-oncology. 2021 Aug 1;23(8):1231-51. 10.1093/neuonc/noab106

3. Jack Jr CR, Bennett DA, Blennow K, Carrillo MC, Dunn B, Haeberlein SB, Holtzman DM, Jagust W, Jessen F, Karlawish J, Liu E. NIALJAA research framework: toward a biological definition of Alzheimer’s disease. Alzheimer’s & dementia. 2018 Apr;14(4):535–62. 10.1016/j.jalz.2018.02.018

4. Mattson MP, Arumugam TV. Hallmarks of brain aging: adaptive and pathological modification by metabolic states. Cell metabolism. 2018 Jun 5;27(6):1176–99. doi:10.1016/j.cmet.2018.05.011

5. Gieryng A, Pszczolkowska D, Walentynowicz KA, Rajan WD, Kaminska B. Immune microenvironment of gliomas. Laboratory investigation. 2017 May 1;97(5):498–518. 10.1038/labinvest.2017.19

6. Quail DF, Joyce JA. The microenvironmental landscape of brain tumors. Cancer cell. 2017 Mar 13;31(3):326–41. doi: 10.1016/j.ccell.2017.02.009

7. Wang Q, Hu B, Hu X, Kim H, Squatrito M, Scarpace L, DeCarvalho AC, Lyu S, Li P, Li Y, Barthel F. Tumor evolution of glioma-intrinsic gene expression subtypes associates with immunological changes in the microenvironment. Cancer cell. 2017 Jul 10;32(1):42–56. doi:10.1016/j.ccell.2017.06.003

8. Colaprico A, Silva TC, Olsen C, Garofano L, Cava C, Garolini D, Sabedot TS, Malta TM, Pagnotta SM, Castiglioni I, Ceccarelli M. TCGAbiolinks: an R/Bioconductor package for integrative analysis of TCGA data. Nucleic acids research. 2016 May 5;44(8):e71-.. doi: 10.1093/nar/gkv1507

9. Therneau TM. Extending the Cox model. InProceedings of the first Seattle symposium in biostatistics: survival analysis 1997 (pp. 51-84). New York, NY: Springer US. doi: 10.1007/978-1-4684-6316-3_5

10. Simon N, Friedman JH, Hastie T, Tibshirani R. Regularization paths for Cox’s proportional hazards model via coordinate descent. Journal of statistical software. 2011 Mar 9;39:1–3. doi: 10.18637/jss.v039.i05

11. Huber M, Bello C, Schober P, Filipovic MG, Luedi MM. Decision curve analysis of in-hospital mortality prediction models: the relative value of pre-and intraoperative data for decision-making. Anesthesia and analgesia. 2024 Feb 5;139(3):617. doi: 10.1213/ANE.0000000000006874

12. Zhao Z, Zhang KN, Wang Q, Li G, Zeng F, Zhang Y, Wu F, Chai R, Wang Z, Zhang C, Zhang W. Chinese Glioma Genome Atlas (CGGA): a comprehensive resource with functional genomic data from Chinese glioma patients. Genomics, proteomics & bioinformatics. 2021 Feb;19(1):1–2. doi: 10.1016/j.gpb.2020.10.005

13. Fjell AM, Walhovd KB, Fennema-Notestine C, McEvoy LK, Hagler DJ, Holland D, Brewer JB, Dale AM. One-year brain atrophy evident in healthy aging. Journal of Neuroscience. 2009 Dec 2;29(48):15223–31. doi: 10.1523/JNEUROSCI.3252-09.2009

14. Neftel C, Laffy J, Filbin MG, Hara T, Shore ME, Rahme GJ, Richman AR, Silverbush D, Shaw ML, Hebert CM, Dewitt J. An integrative model of cellular states, plasticity, and genetics for glioblastoma. Cell. 2019 Aug 8;178(4):835–49. Doi: 10.1016/j.cell.2019.06.024

15. Patel AP, Tirosh I, Trombetta JJ, Shalek AK, Gillespie SM, Wakimoto H, Cahill DP, Nahed BV, Curry WT, Martuza RL, Louis DN. Single-cell RNA-seq highlights intratumoral heterogeneity in primary glioblastoma. Science. 2014 Jun 20;344(6190):1396–401. DOI: 10.1126/science.1254257

16. Verhaak RG, Hoadley KA, Purdom E, Wang V, Qi Y, Wilkerson MD, Miller CR, Ding L, Golub T, Mesirov JP, Alexe G. Integrated genomic analysis identifies clinically relevant subtypes of glioblastoma characterized by abnormalities in PDGFRA, IDH1, EGFR, and NF1. Cancer cell. 2010 Jan 19;17(1):98-110. doi: 10.1016/j.ccr.2009.12.020

17. Van den Bent MJ, Tesileanu CM, Wick W, Sanson M, Brandes AA, Clement PM, Erridge S, Vogelbaum MA, Nowak AK, Baurain JF, Mason WP. Adjuvant and concurrent temozolomide for 1p/19q non-co-deleted anaplastic glioma (CATNON; EORTC study 26053-22054): second interim analysis of a randomised, open-label, phase 3 study. The Lancet Oncology. 2021 Jun 1;22(6):813–23. doi: 10.1016/S1470-2045(21)00090-5

18. Buckner JC, Shaw EG, Pugh SL, Chakravarti A, Gilbert MR, Barger GR, Coons S, Ricci P, Bullard D, Brown PD, Stelzer K. Radiation plus procarbazine, CCNU, and vincristine in low-grade glioma. New England Journal of Medicine. 2016 Apr 7;374(14):1344–55. doi:10.1056/NEJMoa1500925

19. Hickman SE, Kingery ND, Ohsumi TK, Borowsky ML, Wang LC, Means TK, El Khoury J. The microglial sensome revealed by direct RNA sequencing. Nature neuroscience. 2013 Dec;16(12):1896–905. doi:10.1038/nn.3554

20. Stevens B, Allen NJ, Vazquez LE, Howell GR, Christopherson KS, Nouri N, Micheva KD, Mehalow AK, Huberman AD, Stafford B, Sher A. The classical complement cascade mediates CNS synapse elimination. Cell. 2007 Dec 14;131(6):1164–78. doi: 10.1016/j.cell.2007.10.036

21. Colonna M, Wang Y. TREM2 variants: new keys to decipher Alzheimer disease pathogenesis. Nature Reviews Neuroscience. 2016 Apr;17(4):201–7. doi:10.1038/nrn.2016.7

22. Vickers AJ, Cronin AM, Elkin EB, Gonen M. Extensions to decision curve analysis, a novel method for evaluating diagnostic tests, prediction models and molecular markers. BMC medical informatics and decision making. 2008 Nov 26;8(1):53. doi:10.1186/1472-6947-8-53

